# Demonstrating an approach for evaluating synthetic geospatial and temporal epidemiologic data utility: Results from analyzing >1.8 million SARS-CoV-2 tests in the United States National COVID Cohort Collaborative (N3C)

**DOI:** 10.1101/2021.07.06.21259051

**Authors:** Jason A. Thomas, Randi E. Foraker, Noa Zamstein, Philip R.O. Payne, Adam B. Wilcox, the N3C Consortium

## Abstract

**Objective:** To evaluate whether synthetic data derived from a national COVID-19 data set could be used for geospatial and temporal epidemic analyses.

**Materials and Methods:** Using an original data set (n=1,854,968 SARS-CoV-2 tests) and its synthetic derivative, we compared key indicators of COVID-19 community spread through analysis of aggregate and zip-code level epidemic curves, patient characteristics and outcomes, distribution of tests by zip code, and indicator counts stratified by month and zip code. Similarity between the data was statistically and qualitatively evaluated.

**Results:** In general, synthetic data closely matched original data for epidemic curves, patient characteristics, and outcomes. Synthetic data suppressed labels of zip codes with few total tests (mean=2.9±2.4; max=16 tests; 66% reduction of unique zip codes). Epidemic curves and monthly indicator counts were similar between synthetic and original data in a random sample of the most tested (top 1%; n=171) and for all unsuppressed zip codes (n=5,819), respectively. In small sample sizes, synthetic data utility was notably decreased.

**Discussion:** Analyses on the population-level and of densely-tested zip codes (which contained most of the data) were similar between original and synthetically-derived data sets. Analyses of sparsely-tested populations were less similar and had more data suppression.

**Conclusion:** In general, synthetic data were successfully used to analyze geospatial and temporal trends. Analyses using small sample sizes or populations were limited, in part due to purposeful data label suppression -an attribute disclosure countermeasure. Users should consider data fitness for use in these cases.

## INTRODUCTION

### Background and significance

COVID-19 has illustrated the need to disseminate accurate, timely, and useful epidemiologic public health data -especially data related to ongoing pandemics or pandemic preparedness. It has also highlighted the need to protect the privacy of individuals.[1,2] The National COVID Cohort Collaborative (N3C) was created to share and harmonize individual-level electronic health record (EHR) data into a single data set.[3] The N3C has received, ingested, harmonized, and characterized[4] data from across the United States (US). To balance data access and privacy, N3C created two levels of data sets: (1) the limited data set (LDS) which has 16 HIPAA Privacy Rule[5,6] direct identifiers stripped out except dates and zip codes, and (2) synthetic data which are computationally derived from the LDS to mimic the LDS data statistical distributions, covariance, and higher order interactions. Synthetic data generation can potentially protect privacy because synthetic data rows are not directly tied to the original source data.[7–11] Pending a pilot study and privacy validation, synthetic data sets are the only data under consideration to be shared outside of the N3C enclave.[3]

Applying privacy-preserving methods to data comes at varying cost to utility, producing a privacy-utility trade-off.[9,12–15] De-identification removes granular geographic information such as street-level address.

Obscuring dates reduces the utility of temporal data for some analyses, such as epidemic curves. However, these geographic and temporal data are critical components needed to measure key indicators of COVID-19 community spread[16] used to inform pandemic management decisions such as determining when to reopen schools[17] and businesses.[18] Thus, synthetic data are likely the only privacy-preserving N3C data that can be used to analyze some of the most critically-important data related to pandemic management and preparedness while also providing citizens more transparency into the underlying data. However, previous research has reported deficits in how well synthetic data mimic original data including limitations in their: ability to capture longitudinal relationships, model multiple data types, and perform well on small sample sizes[10,19,20]. Due to the combination of potential widespread synthetic data dissemination, heightened research interest in COVID-19[21], and the rise of “citizen science”[22–24], the user base and applications of pandemic-related synthetic data will likely be heterogenous and broad. Therefore, it is important to evaluate N3C synthetic data in a manner that can inform users with a wide range of intended use cases and definitions for synthetic data fitness for use.[25]

The utility of synthetic health data has been evaluated in other work[15,19,20,26–30] outside of N3C which applied a variety of the ways one can validate synthetic data.[31] However, N3C synthetic data utility has only been evaluated once before. Recently, the N3C synthetic data validation task team evaluated the utility of N3C synthetic data (MDClone, Beer Sheva, Israel) across three use cases, one of which had a geospatial and temporal focus.[32] Foraker et al. (2021) found the synthetic data had high utility for construction of a single aggregate epidemic curve of COVID-19 cases. However, it showed that rural zip codes with smaller population counts were more likely than urban zip codes to have zip code labels censored (suppressed) in the synthetic data, which is where a categorical variable’s value is replaced with the word ‘censored’ to protect privacy of patients with particularly uncommon, and thus identifiable, features. To date, no analyses have been conducted on the N3C synthetic data to assess utility for analyses by individual zip codes and/or aggregate indicators beyond case counts (e.g. percent positive) over time.

### Objective

In this paper, we describe the N3C Synthetic data validation task team methods and results focused on evaluating whether synthetic N3C data can be used for geospatial and temporal epidemic analyses. Our replication studies focused on what we deemed were important and common analyses to be performed, such as epidemic curves for key indicators and creation of public-facing dashboards.[33–35] Our validation included replication of studies and general utility metrics[31] for: analyses at the zip code level over time, construction of epidemic curves, and aggregate population characteristics. We believe these approaches balance the need to provide broad utility results for a wide range of analyses while also providing specific validation results relevant to analyses of common interest.

## MATERIALS AND METHODS

### Data

The N3C data analyzed include individual-level EHR data enriched with social determinants of health (SDOH) at the 5-digit zip code level. The data have been harmonized into the Observational Medical Outcomes Partnership (OMOP) common data model (CDM) v5.3.1[3,36] and are the same data sets described in a previous N3C synthetic data validation use case.[32] The N3C LDS as of November 30, 2020 -which included 34 data source partners -was used as the data source. MDClone received a copy of the LDS then transformed these data from the N3C harmonized data model into MDClone’s data model. Afterwards, the required data needed for the study team’s analyses were extracted by MDClone from the transformed LDS for use as the “original” data set. A synthetic derivative of this transformed original data set was then created by MDClone.

MDClone provided both the original and synthetic data sets to the research team for evaluation within the N3C secure enclave environment (flowchart Figure S3 in supplement).

Both the original and synthetic data were formatted as a single table adhering to the same schema, with each row representing a single COVID-19 test. The table had the following columns: test result (positive/negative; only each patient’s first negative and/or first positive test included), age at confirmed test result; admission start date days from reference if admission occurred within ±7 days of COVID-19 positive test result; death (null/yes) during admission; admission length of stay (LOS); patient’s state of residence; source partner with which the patient was affiliated; and patient’s 5-digit zip code. The data also included the following SDOH columns determined by the patient’s zip code: total population in zip code; percent of residents under the poverty line; percent without health insurance; and median household income.

As in Foraker et al. (2021), we used consistent definitions for censored and uncensored zip codes.

Censored zip codes were those present within the original data not found (n=11,222) within the synthetic data set either because the zip code was suppressed by labeling the zip code ‘censored’ or removed within the synthetic data set to protect privacy. Conversely, uncensored zip codes were defined as discrete zip codes found in the original and the synthetic data (n=5,819).

### Analysis

All analyses were conducted solely by one author (JAT). All code was written in Python (v3.6.10) and -as required by N3C -ran within the secure N3C enclave using the Palantir Foundry Analytic Platform (Palantir Technologies, Denver, CO). The entirety of code used in this analysis is contained within a single Foundry Code Workbook using a saved Spark environment to preserve required software versions and dependencies.

The code workbook and source data have been stored within the N3C enclave so that they may inform and be reused in future validation work.

### Summary of data

Descriptive statistics were calculated and reported in Table 1 for age, number of unique zip codes present, LOS, and admission date after positive test stratified by patients who were tested, positive, admitted, and who died during admission. Number of unique zip codes present excluded null or censored zip codes. The difference between original and synthetic values was reported as the raw synthetic difference (synthetic -original). The difference as a percentage of the original value was reported as synthetic difference percentage (raw synthetic difference/original).

**Table 1.** Testing and outcomes characteristics: comparison of original vs synthetic data

|  | Original | Synthetic | Synthetic difference (raw) | Synthetic difference (%) |
| --- | --- | --- | --- | --- |
| **Tests (n)** | 1854968 | 1854950 | -18.00 | 0.00 |
| Age (mean) | 44 | 44 | 0.00 | 0.00 |
| Age (stdev) | 22.16 | 22.16 | 0.00 | 0.00 |
| Age (median) | 43.52 | 43.51 | -0.01 | -0.02 |
| Age (IQR) | 35.08 | 35.04 | -0.04 | -0.11 |
| Unique zip codes (n) | 17041 | 5819 | -11222.00 | -65.85 |
| **Positive (count)** | 195200 | 195198 | -2.00 | 0.00 |
| Positive (%) | 10.52 | 10.52 | 0.00 | 0.00 |
| Age (mean) | 41.54 | 41.53 | -0.01 | -0.02 |
| Age (stdev) | 20.4 | 20.42 | 0.02 | 0.10 |
| Age (median) | 39.65 | 39.56 | -0.09 | -0.23 |
| Age (IQR) | 31.84 | 31.81 | -0.03 | -0.09 |
| Unique zip codes (n) | 6660 | 1798 | -4862.00 | -73.00 |
| **Negative (n)** | 1659768 | 1659752 | -16.00 | 0.00 |
| Negative (%) | 89.48 | 89.48 | 0.00 | 0.00 |
| Age (mean) | 44.29 | 44.29 | 0.00 | 0.00 |
| Age (stdev) | 22.34 | 22.34 | 0.00 | 0.00 |
| Age (median) | 44.08 | 44.08 | 0.00 | 0.00 |
| Age (IQR) | 35.36 | 35.34 | -0.02 | -0.06 |
| Unique zip codes (n) | 16668 | 5805 | -10863.00 | -65.17 |
| **Admitted (n)** | 23044 | 23044 | 0.00 | 0.00 |
| Admitted (%) | 1.24 | 1.24 | 0.00 | 0.00 |
| Age (mean) | 57.87 | 57.85 | -0.02 | -0.03 |
| Age (stdev) | 19.77 | 19.74 | -0.03 | -0.15 |
| Age (median) | 59.98 | 60 | 0.02 | 0.03 |
| Age (IQR) | 28.2 | 28.22 | 0.02 | 0.07 |
| Days after positive test (mean) | -0.07 | -0.1 | -0.03 | 42.86 |
| Days after positive test (stdev) | 1.77 | 1.74 | -0.03 | -1.69 |
| Days after positive test (median) | -0.05 | -0.04 | 0.01 | -20.00 |
| Days after positive test (iqr) | 0.88 | 0.88 | 0.00 | 0.00 |
| LOS (mean) | 6.48 | 8.32 | 1.84 | 28.40 |
| LOS (stdev) | 290.81 | 10.66 | -280.15 | -96.33 |
| LOS (median) | 5 | 5 | 0.00 | 0.00 |
| LOS (IQR) | 8 | 8 | 0.00 | 0.00 |
| Unique zip codes (n) | 3132 | 1515 | -1617.00 | -51.63 |
| **Died (n)** | 2032 | 2032 | 0.00 | 0.00 |
| Died (%) | 0.11 | 0.11 | 0.00 | 0.00 |
| Age (mean) | 71.81 | 71.81 | 0.00 | 0.00 |
| Age (stdev) | 14.57 | 14.65 | 0.08 | 0.55 |
| Age (median) | 73.26 | 73.21 | -0.05 | -0.07 |
| Age (IQR) | 19.68 | 19.58 | -0.10 | -0.51 |
| Days after positive test (mean) | -0.32 | -0.32 | 0.00 | 0.00 |
| Days after positive test (stdev) | 1.39 | 1.36 | -0.03 | -2.16 |
| Days after positive test (median) | -0.14 | -0.11 | 0.03 | -21.43 |
| Days after positive test (IQR) | 0.91 | 0.93 | 0.02 | 2.20 |
| LOS (mean) | 13.69 | 13.71 | 0.02 | 0.15 |
| LOS (stdev) | 12.93 | 13.05 | 0.12 | 0.93 |
| LOS (median) | 10 | 10 | 0.00 | 0.00 |
| LOS (IQR) | 13 | 13 | 0.00 | 0.00 |
| Unique zip codes (n) | 831 | 16 | -815.00 | -98.07 |

### Aggregate epidemic curves

We constructed aggregate epidemic curves using each data set spanning January 1st through November 30th 2020. The following key indicators were calculated and visualized: tests, cases (reproduced from Foraker et al., 2021, to view others in context), percent positive, admissions, and deaths during admission. Each indicator had the following daily metrics calculated: count (discrete indicators) or value (continuous indicators), 7-day midpoint moving average, 7-day slope (count or daily value -its value six days prior). To assess the statistical difference between original and synthetic epidemic curves, we conducted the paired two-sided t-test (scipy v1.5.3, stats.ttest_rel) and two-sided wilcoxon signed-rank test (scipy v1.5.3, stats.wilcoxon) for all metrics across all indicators (Table 2), treating each data set’s daily results as a pair.

**Table 2.** Tests for significant differences between aggregate original and synthetic epidemic curves.

| Key Indicator | Metric | Wilcoxon result | P-value | T-test stat | P-value |
| --- | --- | --- | --- | --- | --- |
| Tests | Counts | 25354.5 | 0.300 | -0.007 | 0.994 |
|  | 7-day average | 25458.5 | 0.428 | -0.025 | 0.980 |
|  | 7-day slope | 26075 | 0.735 | -0.002 | 0.998 |
| Cases | Counts | 26288 | 0.496 | -0.002 | 0.998 |
|  | 7-day average | 26005 | 0.775 | -0.006 | 0.996 |
|  | 7-day slope | 25788.5 | 0.898 | -0.002 | 0.998 |
| Percent Positive | Counts | 26407 | 0.426 | -0.932 | 0.352 |
|  | 7-day average | 24038 | 0.072 | -2.258 | **0.025** |
|  | 7-day slope | 27083 | 0.972 | 0.129 | 0.896 |
| Admissions | Counts | 21405 | 0.247 | -0.007 | 0.995 |
|  | 7-day average | 24299 | 0.197 | -0.030 | 0.976 |
|  | 7-day slope | 22825.5 | 0.894 | -0.011 | 0.991 |
| Deaths | Counts | 13881 | 0.748 | 0 | 1 |
|  | 7-day average | 19171.5 | 0.247 | -0.023 | 0.982 |
|  | 7-day slope | 16632 | 0.866 | -0.011 | 0.992 |

### Distribution of tests; censoring of zip codes

To assess the distribution of tests by zip code and threshold of zip code censoring, we calculated the total number of tests per zip code in the original and synthetic data. In the synthetic data, we excluded rows with a censored (n=44,337; 2.4%) or null (n=444,092; 23.9%) zip code. In the original data, we excluded rows with a null (n=444,380; 24.0%) zip code. We computed the 99th, 97.5th, and 90th percentiles of tests per zip code in the original data. The distributions of tests by zip code were plotted as a histogram (Figure 2) with the synthetic and original data overlaid. Additionally, we calculated the distribution of tests by zip code in the original data that were censored in the synthetic data, then plotted the result as a histogram (Figure S2 in supplement). We then calculated the difference in patients’ SDOH values within the original data, comparing patients whose zip codes were censored within the synthetic data to those whose zip codes were not censored (Table 3).

**Figure 1:**
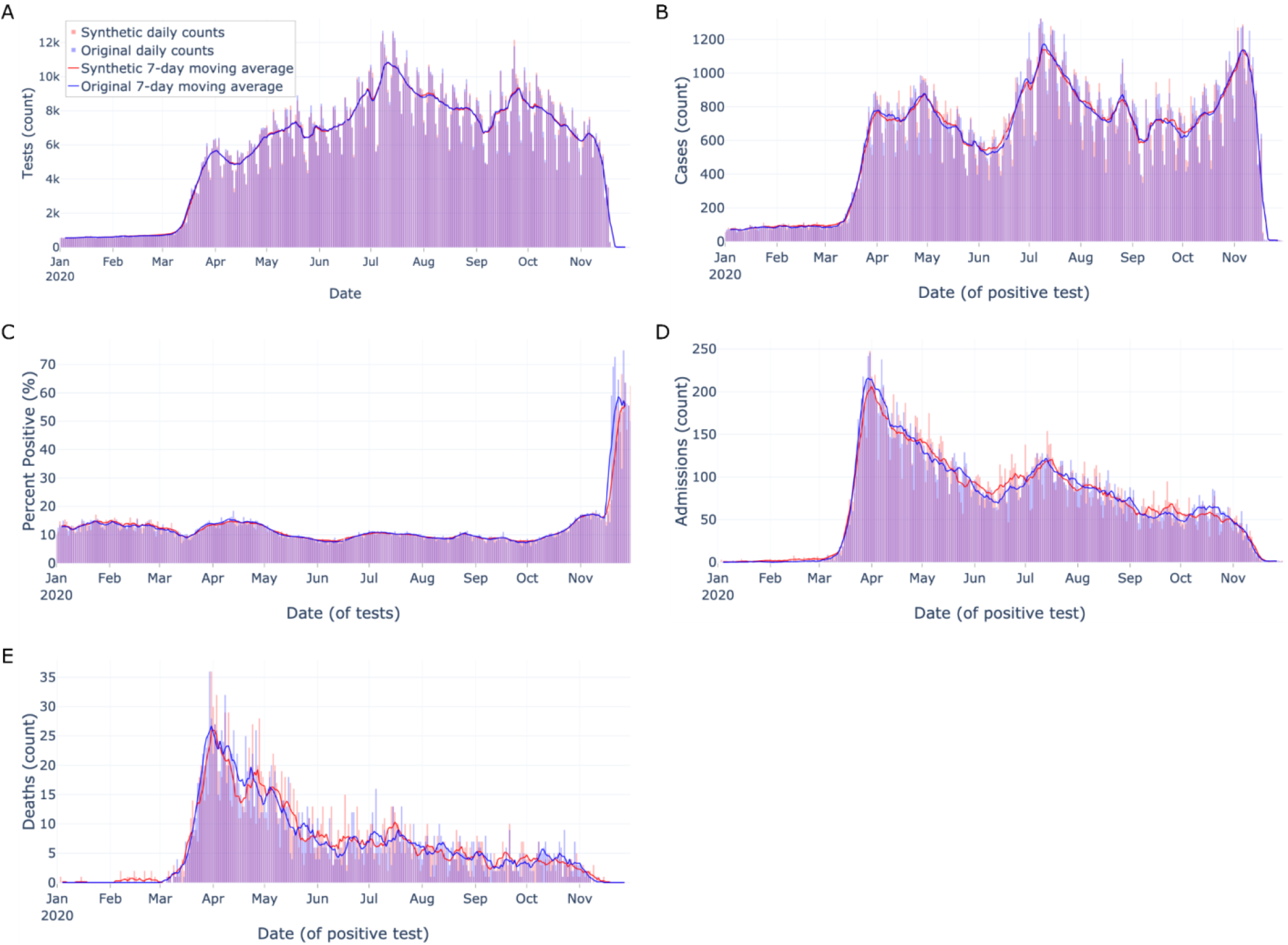
Aggregate epidemic curves of key indicators Aggregate epidemic curves with counts (vertical bars) and 7-day moving averages (smoothed line) for A) tests, B) cases, C) percent positive, D) admissions, and E) deaths during admission. Color encodings include original data (light blue) and synthetic data (light red), with their overlap (purple). As counts get smaller from tests to deaths, the epidemic curves visually appear less similar.

**Figure 2:**
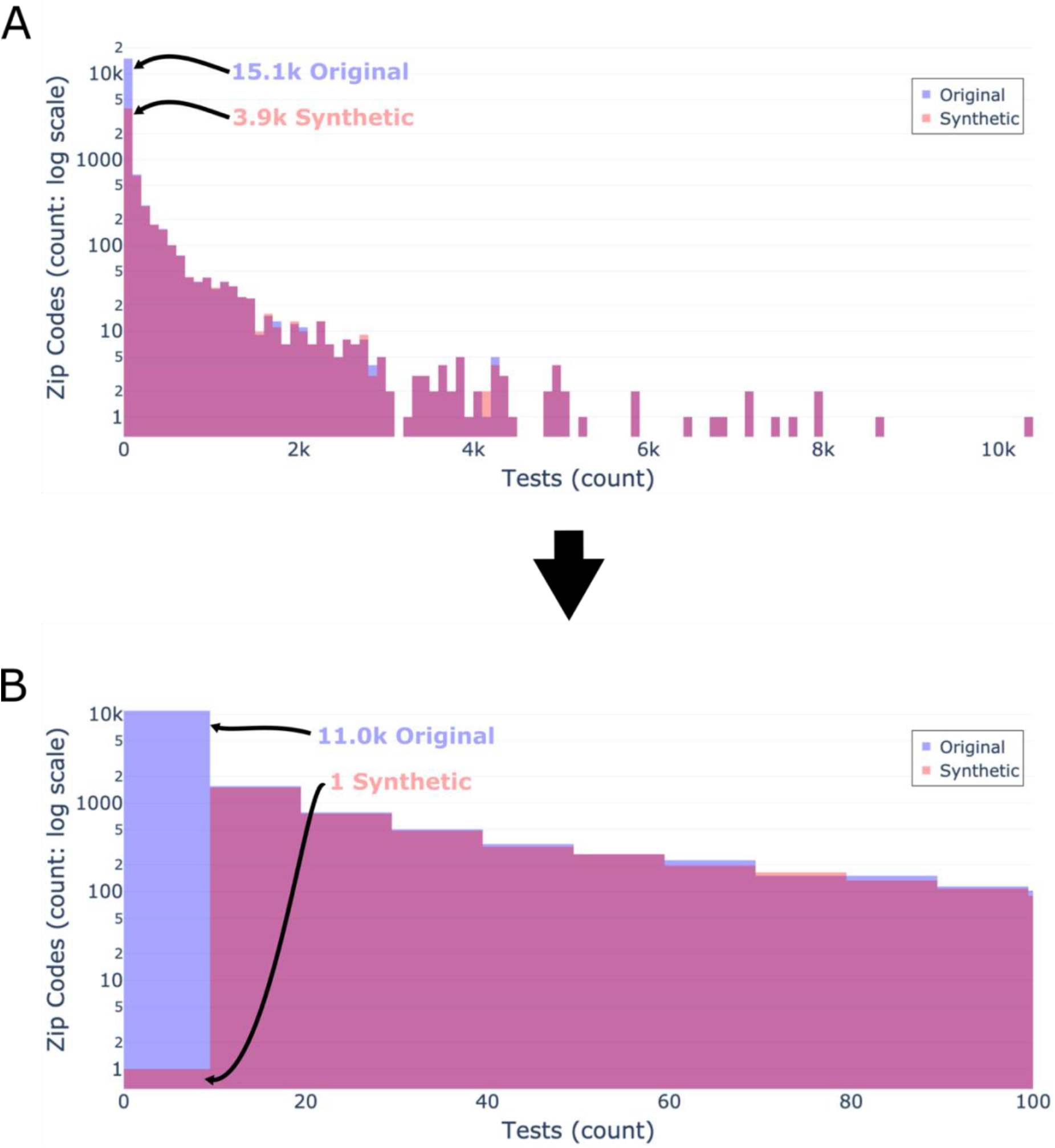
Distribution of tests by zip code Distributions of total tests by zip code shown by original data (light blue) and synthetic data (light red), and their overlap (purple). A) All data binned by 100. B) Filtered data with a bin size of 10 to only show the distribution of tests by zip code in zip codes with <100 tests. Both y-axes use a log scale. As seen in panel A, the vast majority of tests are conducted in a minority of zip codes. As seen in panels A & B, the distribution of the synthetic data closely matches the original data at >10 tests per zip code.

**Table 3.** SDOH and age of patients in the original data whose zip codes were censored vs. uncensored

| SDOH | Censored Status | Mean | Standard deviation | Median | IQR |
| --- | --- | --- | --- | --- | --- |
| Age (years) | Uncensored | 44.0 | 22.2 | 43.5 | 35.0 |
|  | Censored | 46.4 | 22.0 | 48.7 | 40.1 |
|  | Uncensored Difference (raw) | -2.4 | 0.2 | -5.2 | -5.1 |
| Median household income (\$) | Uncensored | 64092.6 | 23973.9 | 59324.0 | 29241.0 |
|  | Censored | 63101.5 | 28964.1 | 55625.0 | 28857.0 |
|  | Uncensored Difference (raw) | 991.1 | -4990.2 | 3699.0 | 384.0 |
| Percent under the poverty line | Uncensored | 13.7 | 9.0 | 11.3 | 11.2 |
|  | Censored | 13.3 | 9.6 | 11.2 | 10.9 |
|  | Uncensored Difference (raw) | 0.4 | -0.6 | 0.1 | 0.3 |
| Percent without health insurance | Uncensored | 8.7 | 5.1 | 7.6 | 7.0 |
|  | Censored (raw) | 9.2 | 6.7 | 7.8 | 7.7 |
|  | Uncensored Difference (raw) | -0.5 | -1.6 | -0.2 | -0.7 |
| Total population of zip code | Uncensored | 29758.7 | 17992.4 | 28479.0 | 25220.0 |
|  | Censored | 15493.9 | 17967.1 | 7935.0 | 23119.3 |
|  | Uncensored Difference (raw) | 14264.8 | 25.3 | 20544.0 | 2100.7 |

### Top 1% paired zip codes’ epidemic curves

Next, we assessed synthetic epidemic curves’ performance at the zip code level, focusing on zip codes with relatively abundant data. We created a list of zip codes from the original data in the 99th percentile (n=171) by total number of tests, then removed any zip codes without an uncensored matched zip code pair in the synthetic data (n=0). We randomly sampled ten zip codes from the list and constructed epidemic curves for these zip codes’ original and synthetic data (Figures 3-4). Each epidemic curve was constructed using the same date range, methods, and metrics as the aggregate epidemic curves described above with the following change: we only assessed tests and admissions indicators due to the infrequency of death during admission at the zip code level and manuscript space limitations.

**Figure 3:**
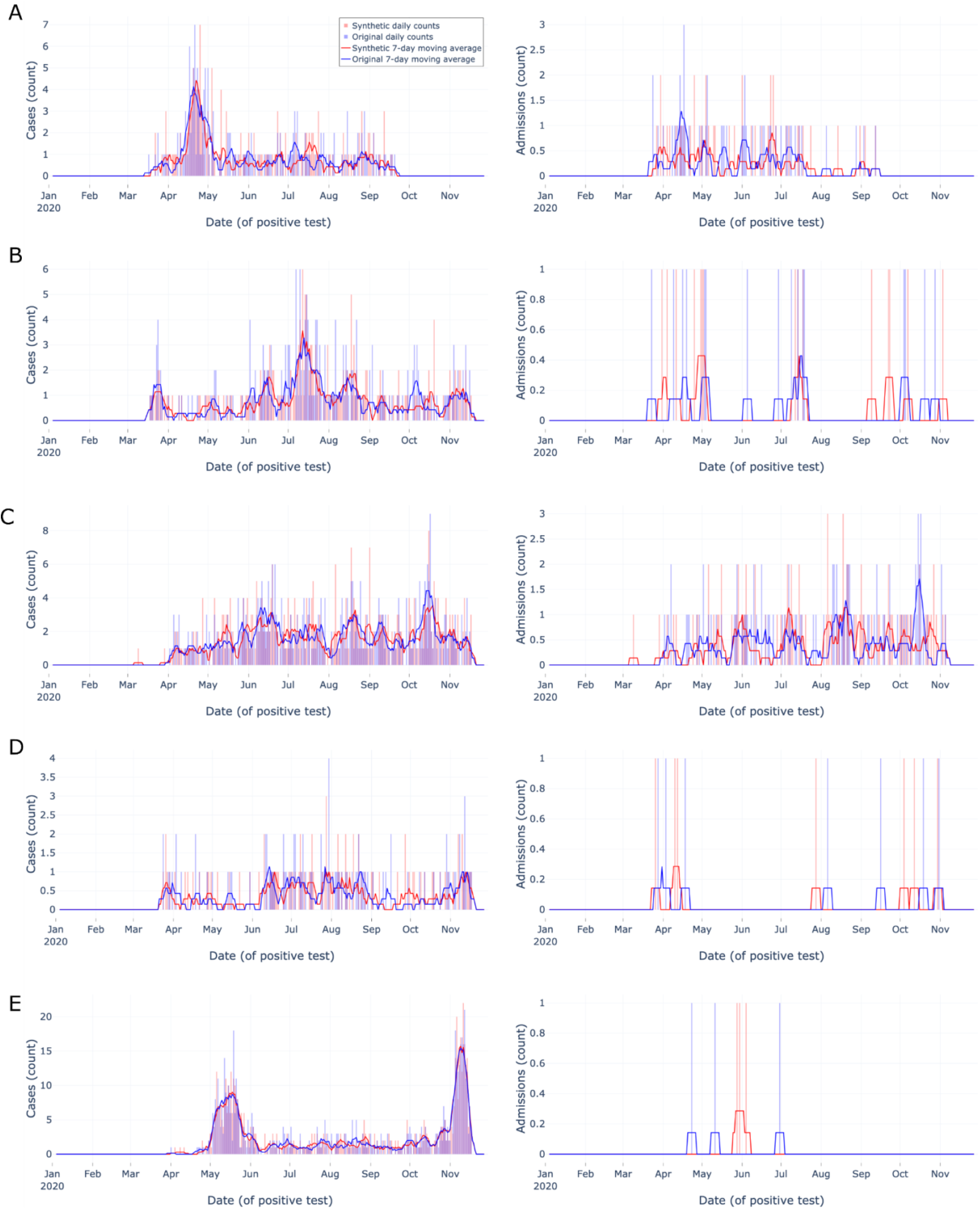
Zip code-level epidemic curves for random sample set #1 of the most tested (top 1%) zip codes Zip code-level epidemic curves with counts (vertical bars) and 7-day moving averages (smoothed line). Color encodings include original data (light blue) and synthetic data (light red), with their overlap (purple). Each row (A-E) corresponds to a different randomly sampled zip code visualizing cases (left column) and admissions (right column). Synthetic data are more similar to original data when indicator density is higher, Overall, synthetic data closely match overall trends and closely match start and end dates.

**Figure 4:**
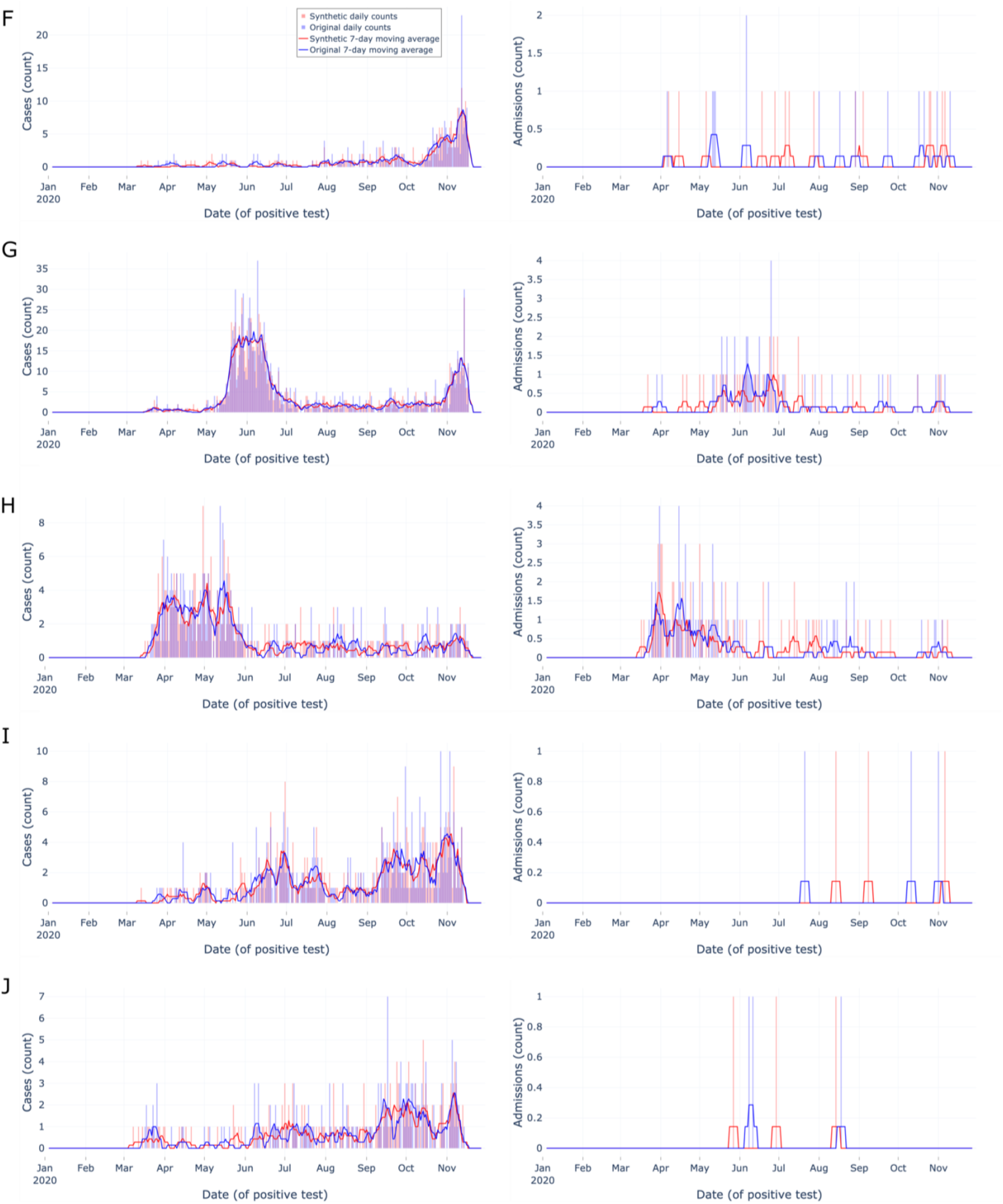
Zip code-level epidemic curves for random sample set #2 of the most tested (top 1%) zip codes Zip code-level epidemic curves with counts (vertical bars) and 7-day moving averages (smoothed line). Color encodings include original data (light blue) and synthetic data (light red), with their overlap (purple). Each row (F-J) corresponds to a different randomly sampled zip code visualizing cases (left column) and admissions (right column). Synthetic data are more similar to original data when indicator density is higher, Overall, synthetic data closely match overall trends and closely match start and end dates.

### Monthly zip code pairwise synthetic error

We compared the difference in monthly counts of tests, cases, and admissions between the original data and paired uncensored synthetic zip codes. To do so, we calculated each data set’s number of tests, cases and admissions for every zip code stratified by month for each month the zip code had ≥ 1 test. Then, the data sets were outer merged on month and zip code (Figure 5). Synthetic error, defined as the difference between the synthetic monthly count and the original data monthly count value, was computed for every zip code month pair. The distribution of synthetic error was visualized (Figure 6) for tests, cases, and admissions.

**Figure 5:**
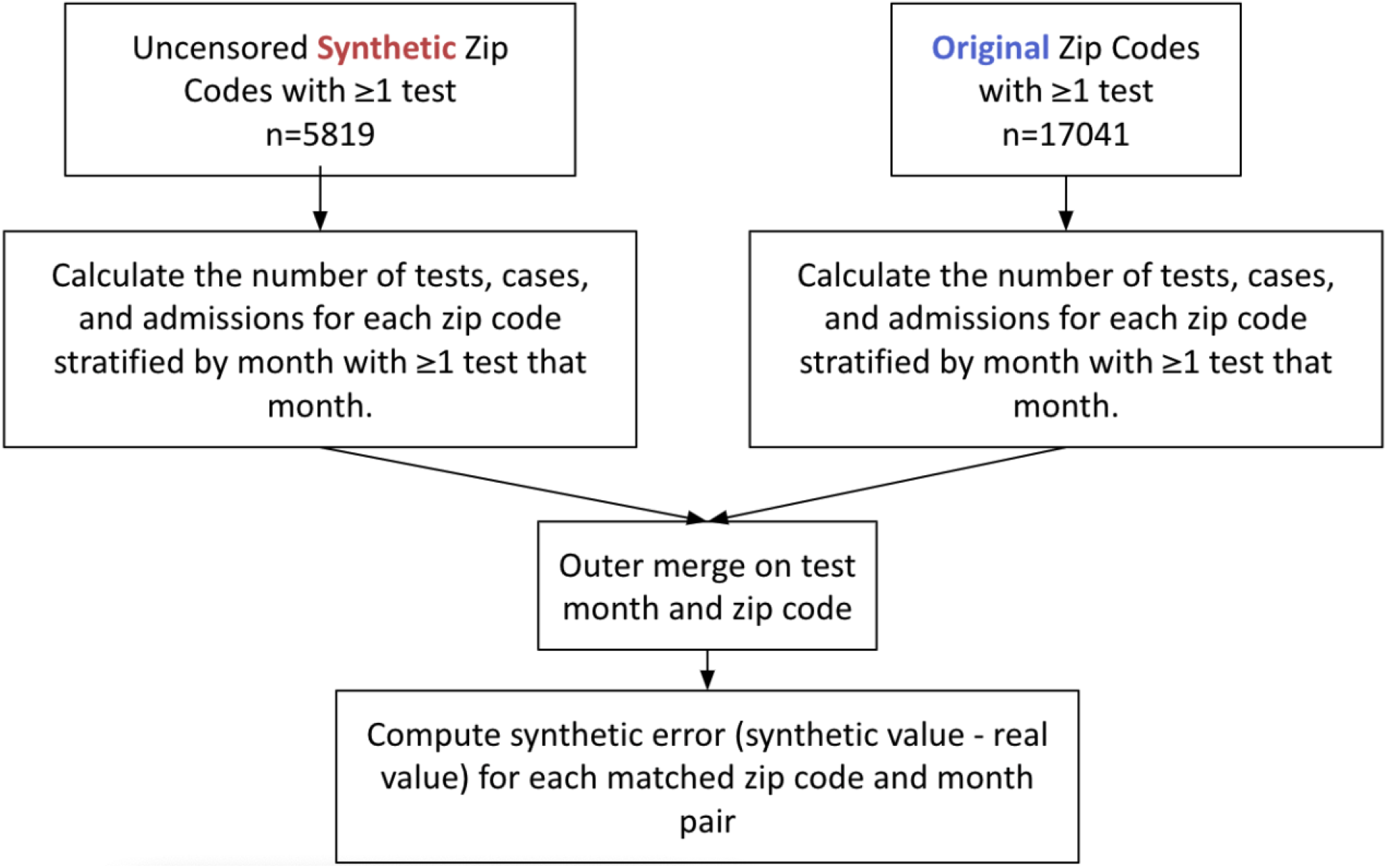
Workflow of synthetic error stratified by zip code and month analysis Workflow of synthetic error experiment showing synthetic data on the left, original data on the right which are then merged to allow the calculation of synthetic error to be made.

**Figure 6:**
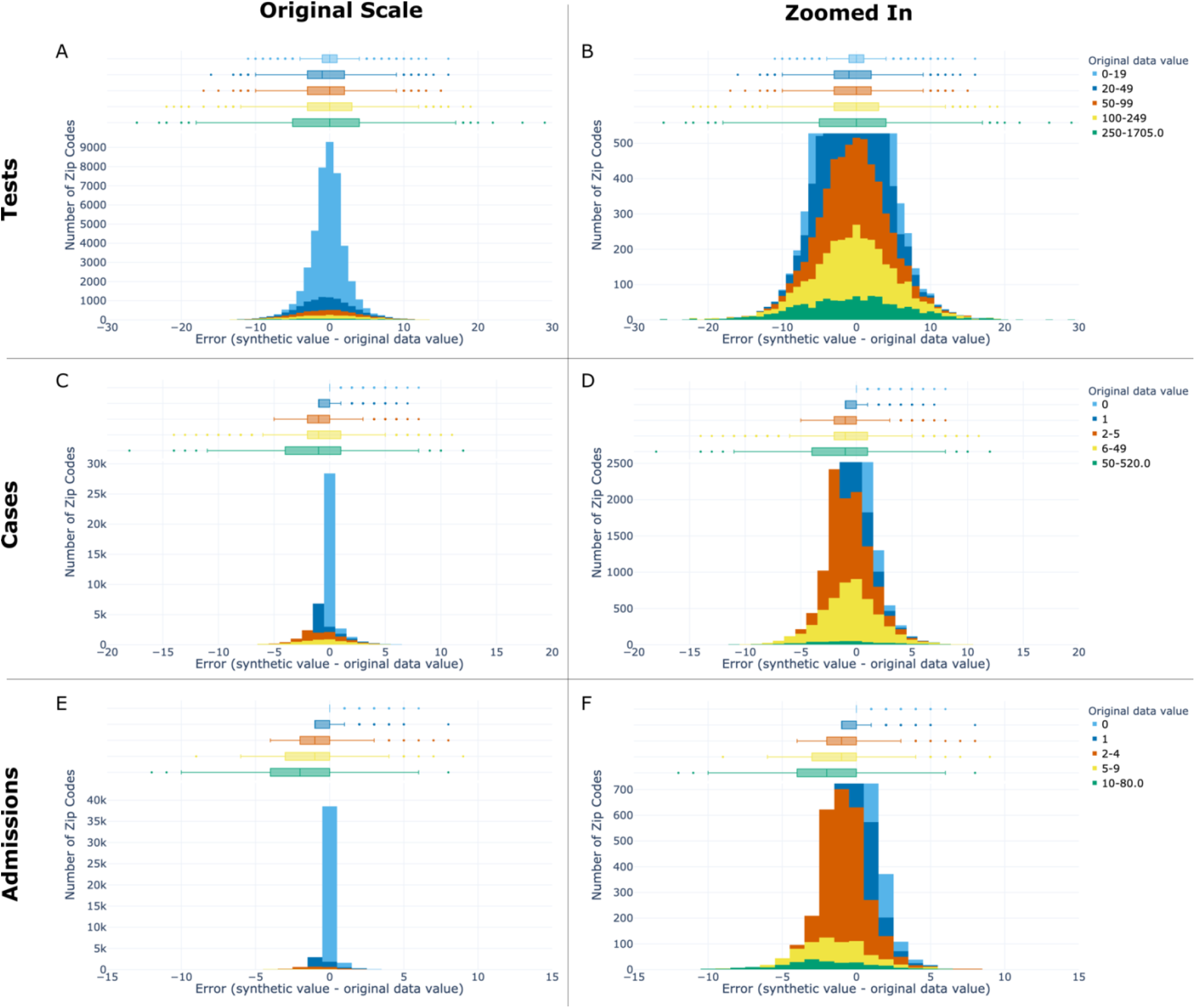
Synthetic error stratified by zip code and month Synthetic error distributions per zip code stratified by month for tests (top row), cases (middle row), and admissions (bottom row) shown both at original scale (left column) and zoomed in to the peak of each row’s middle bin (legend showing bin ranges and color encodings seen on the far right of each row). Original data value denotes the monthly count in the original data for the key indicator of interest. Box plots of synthetic error are shown in the top 30% of each sub-plot (A-F), with a histogram of synthetic error shown in the bottom 70%. Within each sub-plot, the box plot and histogram have a shared x-axis corresponding to synthetic error and shared bins corresponding to the original data value. The y-axis shows the number of zip codes stratified by month (e.g. zip code month pairs). Boxes in the box plots span from Q1 to Q3, with median marked inside the box. Fences span +/-1.5 times the IQR. Error increased as the size (count) of the original data increased, which allows users to estimate the level of error in their data of interest. The synthetic data systematically underestimate the monthly count of key indicators in zip codes with the most tests, cases, and deaths, and overestimate them in zip codes with the least.

### Visualizations

All visualizations (Plotly v4.14.1, Plotly Technologies Inc.) were interactive, allowing N3C enclave users to zoom in/out, pan, and hover to see values and/or labels. In this manuscript, static figures are presented. Log scales were avoided when possible and, when used, annotated to draw attention to the scale.

Visualizations that overlaid both datasets adhered to consistent style conventions. We encoded synthetic and original data sources as red and blue, respectively. Vertical overlaid bars were set to an opacity of 0.35 to 1) provide contrast between two datasets and 2) allow additional tracings, such as 100% opacity 7-day moving averages used in epidemic curves, to be seen on top of the bars.

All visualizations were created using colorblind-safe color mappings. Categorical mappings encoding values besides data source (synthetic or original) used hexadecimal color codes found in the seaborn colorblind palette[37,38]. Each visualization was qualitatively tested for colorblind deuteranopia, protanopia, and tritanopia interpretability by one member of the research team (JAT) using Color Oracle.[39]

## RESULTS

There were nearly two million tested patients (Original n=1,854,968; Synthetic n=1,854,950) in each data set. As seen in Table 1, the overall central tendencies of variables of interest overall were similar between the synthetic data and original data, especially for age and percent positive/admitted/died. The raw synthetic difference was zero, rounded to two decimal points, roughly one third (18/50 rows in Table 1) of the time. The variable with the greatest synthetic difference was unique zip codes, with between a 65-98% reduction in unique zip codes. Median LOS and IQR for admitted patients were exactly the same, yet the mean LOS was 6.48 (±290.81) and 8.32 (±10.66) days for original and synthetic values, respectively. The extreme LOS standard deviation observed in the original data was due to an erroneous outlier. A single row in the original data had an extreme negative LOS [∼-44,000 days; ∼-120 years] and 11 rows with a LOS=-1. The synthetic data also had negative LOS values (n<10) but the values were greatly attenuated, ranging from -1 to roughly -175.

In our statistical analysis, no differences were found between the aggregate epidemic curves besides the 7-day average of percent positive [(t-test p-value=0.025; wilcoxon p-value=0.072), Table 2].

Differences were observed between patients’ SDOH values whose zip codes were uncensored in the synthetic data compared to patients whose zip codes were censored in the synthetic data (Table 3). The largest differences were found in the total population of zip code and age. Patients with uncensored zip codes lived in more populous zip codes (median median total population: uncensored=28,479, censored=7,935) and were younger (median age: uncensored=43.5, censored=48.7).

### Distribution of tests by zip code and of censored zip codes

The 90th, 97.5th, and 99th percentiles for total tests by zip code in the original data were 125, 784, and 1,636 tests, respectively (see Figure 2A). Thus, a small minority of zip codes account for the vast majority of total tests. There were 15,108 (88.7%) unique zip codes in the original data with <100 total tests and 11,039 (64.7%) with <10 tests. Above this threshold (n≥10 tests), the synthetic data mimic the original data distribution closely (see Figure 2B). There were 17,041 unique zip codes and 5,819 unique uncensored zip codes in the original and synthetic data, respectively. The vast majority of censored zip codes are those that had <10 total tests in the original data (mean=2.9±2.4; median=2, IQR=3; max=16) as seen in Figure S2 of the supplement.

### Monthly zip code pairwise synthetic error

The absolute value of pairwise synthetic error stratified by month and zip code increased as the original data value of counts increased (see Figure 6; supplement Table S1). Thus, as sample size of data increased, so did the absolute synthetic error and vice versa. The synthetic error for tests ranged from an IQR=2 when the original value of tests was between 0 to 19 to IQR=9 when the original value of tests was between 250 and 1,705. All synthetic error for zip codes with an original bin value of zero count was positive. All other bins’ synthetic error across key indicators was skewed negative, indicating that the synthetic data had lower counts than the original data.

## DISCUSSION

Overall, analyses on the population-level and of densely-tested zip codes (which contained most of the data) were similar between original and synthetically-derived data sets. Analyses of sparsely-tested populations with smaller sample sizes were notably less similar and had more data suppression, which is in agreement with prior work.[19,32] Synthetic data most closely matched the original data on aggregate data tasks such as aggregate epidemic curves (Figure 1) and broad summary statistics (Table 1). At the aggregate level, only one metric (percent positive, 7-day average) across all indicators showed a significant difference between synthetic and original data aggregate epidemic curves (Table 2). Scarcity of data -as data collection used in this manuscript tapered off in November -is likely a contributing factor to the difference.

The summary statistics shown of both data sets’ populations in Table 1 were similar. Major exceptions were the number of unique zip codes due to censoring in the synthetic data and attenuation in the synthetic data of a single extreme outlier (∼-44,000 day LOS) caused by a data quality issue in the original data. Other erroneous negative LOS values persisted within the synthetic data, yet the bulk of the erroneous values remaining were a LOS=-1 which has been reported as a data quality issue attributed to daylight savings.[40,41] Thus, we show that synthetic data can reduce the impact of data quality issues by removing or attenuating erroneous outliers in order to protect the privacy of rare, and thus identifiable, data.

At the zip code and month level, the synthetic data error performed well on an absolute level; the error increased as the size of the original data increased (Figure 5 & Supplementary Table 1). Therefore, the amount of synthetic error is predictable which gives users the ability to estimate the level of error in their data of interest. Additionally, the synthetic error relative to the original data value is likely small enough for most uses of synthetic data. For example, a zip code in the synthetic data with a monthly positive count of 6-49 is off from the original data by an average of -0.59±2.63. The overrepresentation of negative tests in the original data by 8.5-fold (Table 1) appears to bias synthetic error. Since it is impossible to have less than zero count, the synthetic data cannot add privacy-producing noise in the negative direction for zip code monthly counts equal to zero. Consequently, the synthetic data systematically underestimate the monthly count of key indicators in zip codes with the most tests, cases, and deaths, and overestimate them in zip codes with the least. Our results relate to Flaxman et al., 2020 which observed a similar effect resulting from a non-negativity constraint in the US Census’ TopDown differential privacy algorithm.[12] The magnitude of the synthetic error skewing negative in a smaller concentration of zip codes increased as a key indicator became less frequent, which is fundamentally a signal problem in low-density data sets and is not specific to synthetic data generation.

The top 1% most tested zip codes’ epidemic curves (Figure 3 & Figure 4) provide users with 10 qualitative examples of densely tested zip codes. Overall, the synthetic data closely matched the start and end dates of the original data and followed the overall trend of the original data over time (e.g. Figure 3A matched spike in late April). The ten examples show users the 99th percentile best-case scenario of key indicator original data availability and synthetic data performance at the zip code level, yet the size and testing density of N3C data will likely continue to increase.

Our findings show the importance of understanding the characteristics and limitations of the original data since we found these biases affected synthetic data utility. Data biases resulting in poorer performance of software tools, clinical guidelines and other applications for groups underrepresented in source data has been previously reported for separate tasks.[12,42–45] Foraker et al. (2021) found that censored zip codes had greater missingness of SDOH values in the original data than uncensored zip codes. In our study, we found the bulk of patients in the N3C data live in a small minority of zip codes (Figure 2), likely those most adjacent to institutions contributing data. These zip codes are therefore more likely to be urban and less likely to have their zip code censored (Table 3). As a consequence, rural zip codes, which are already underrepresented in the original data, become even less available to directly analyze. Additionally, patients with censored zip codes were older, potentially due to older patients traveling from sparsely tested regions to receive care offered at distant academic medical centers which participate in N3C.

While our results demonstrate the utility of using synthetic data for a broad range of geospatial analyses, a caveat to synthetic data use is its utility to analyze rural N3C populations since nearly all zip codes with <10 tests were censored and much more likely to be rural within the original data. Suppression of non-zero counts <10 is a common convention within state and federal guidelines to avoid inadvertent disclosure of protected health information for publicly released data.[46–48] Analyses such as choropleth maps at the zip code level including sparsely tested regions would benefit from using the LDS to obtain access to all zip codes without suppression, or by generating and using a different MDClone synthetic dataset that reports geospatial data at a lower level of granularity (e.g. 3-digit zip codes). Our results may inform future N3C discussions about data set balancing ranging from 1) creation of artificially balanced hybrid data sets to improve statistical models’ performance on underrepresented data[42,49], 2) source partners sending a random sample of negative tests alongside all positive tests, or 3) expansion of data ingestion from rural regions.

Whether these synthetic data are “good enough” hinges on a fitness for use determination to be made by each user. The authors believe the data will be useful enough for a wide variety of use cases. Educational software engineering projects or pandemic preparedness tool development could be especially well-served by these data. A major limitation of the data, however, is that they are output in a different data model than the OMOP CDM.[36] Thus, tools built on the synthetic data would not be transferable to run on the LDS without modification. Other users may find the synthetic data well suited to rapid, iterative hypothesis generation/testing without the delays of acquiring the relatively more restricted LDS.[3]

## Limitations and future work

To date, no privacy analysis has been published on these synthetic data to provide context for its utility in relation to its privacy. The data used in this manuscript do not reflect the current size nor state of the N3C LDS. Other statistical techniques such as equivalence testing, bhattacharyya distance[50,51], or adversarial challenges[28] could be used in the future to compare similarity between epidemic curves. The Wilcoxon signed-rank and paired t-tests assume the null hypothesis that the original and synthetic datasets are equivalent. Equivalence testing, which flips the null hypothesis, may be better suited. Equivalence testing was not used in this manuscript due to the challenge of selecting an equivalence bound without knowing what threshold(s) data end-users would find most applicable. Future work conducting equivalence testing specific to well-defined, high-impact use cases may be merited. However, the work required to do so in an ad hoc manner may suggest the LDS is a better alternative in those cases.

## CONCLUSION

Overall, the synthetic data are promising for a wide range of use cases including: population level summary statistics, epidemic curves for the data in aggregate and for the most densely tested zip codes, and analyses necessitating monthly counts of key indicators for the top third of zip codes by number of tests.

However, analyses requiring unsuppressed zip code analyses on populations with <10 tests may be better served by the LDS. Biases found in the original data -namely an underrepresentation of positive tests and tests in rural zip codes -were reflected in the synthetic data. Therefore, it is important to understand the limitations and biases of the original data in addition to the synthetic data impacted downstream from it. We expect the user base of N3C synthetic data to be heterogeneous and the use cases of the data to be broad, resulting in a wide range of fitness for use definitions. To date, there is no published evaluation that quantifies the privacy afforded by this synthetic dataset specifically -nor of the MDClone system itself broadly -to contextualize this synthetic dataset’s utility in relation to a privacy-utility tradeoff; such evaluations are beyond the scope of this work.

Future privacy evaluations of MDClone will not necessarily reflect the privacy of the synthetic data analyzed in this study unless the same dataset and/or the same MDClone system version and parameters are evaluated. Our evaluation of the N3C synthetic data utility provides users the ability to assess whether the synthetic data are fit for use through its combination of general-purpose data utility assessments and visualized replications of analyses of common interest.

## Data Availability

The entirety of code used in this analysis is contained within a single Palantir Foundry Code Workbook using a saved Spark environment to preserve required software versions and dependencies. The code workbook and source data have been stored within the National Covid Cohort Collaborative (N3C) enclave (https://covid.cd2h.org/enclave) so that they may inform and be reused in future validation work. To view National Covid Cohort Collaborative (N3C) Data Enclave & Data Access Requirements, please navigate to the N3C website.
Data was provided from the following institutions: Stony Brook University - U24TR002306, University of Oklahoma Health Sciences Center - U54GM104938: Oklahoma Clinical and Translational Science Institute (OCTSI), West Virginia University - U54GM104942: West Virginia Clinical and Translational Science Institute (WVCTSI), University of Mississippi Medical Center - U54GM115428: Mississippi Center for Clinical and Translational Research (CCTR), University of Nebraska Medical Center - U54GM115458: Great Plains IDeA-Clinical & Translational Research, Maine Medical Center - U54GM115516: Northern New England Clinical & Translational Research (NNE-CTR) Network, Wake Forest University Health Sciences - UL1TR001420: Wake Forest Clinical and Translational Science Institute, Northwestern University at Chicago - UL1TR001422: Northwestern University Clinical and Translational Science Institute (NUCATS), University of Cincinnati - UL1TR001425: Center for Clinical and Translational Science and Training, The University of Texas Medical Branch at Galveston - UL1TR001439: The Institute for Translational Sciences, Medical University of South Carolina - UL1TR001450: South Carolina Clinical & Translational Research Institute (SCTR), University of Massachusetts Medical School Worcester - UL1TR001453: The UMass Center for Clinical and Translational Science (UMCCTS), University of Southern California - UL1TR001855: The Southern California Clinical and Translational Science Institute (SC CTSI), Columbia University Irving Medical Center - UL1TR001873: Irving Institute for Clinical and Translational Research, George Washington Children's Research Institute - UL1TR001876: Clinical and Translational Science Institute at Children's National (CTSA-CN), University of Kentucky - UL1TR001998: UK Center for Clinical and Translational Science, University of Rochester - UL1TR002001: UR Clinical & Translational Science Institute, University of Illinois at Chicago - UL1TR002003: UIC Center for Clinical and Translational Science, Penn State Health Milton S. Hershey Medical Center - UL1TR002014: Penn State Clinical and Translational Science Institute, The University of Michigan at Ann Arbor - UL1TR002240: Michigan Institute for Clinical and Health Research, Vanderbilt University Medical Center - UL1TR002243: Vanderbilt Institute for Clinical and Translational Research, University of Washington - UL1TR002319: Institute of Translational Health Sciences, Washington University in St. Louis - UL1TR002345: Institute of Clinical and Translational Sciences, Oregon Health & Science University - UL1TR002369: Oregon Clinical and Translational Research Institute, University of Wisconsin-Madison - UL1TR002373: UW Institute for Clinical and Translational Research, Rush University Medical Center - UL1TR002389: The Institute for Translational Medicine (ITM), The University of Chicago - UL1TR002389: The Institute for Translational Medicine (ITM), University of North Carolina at Chapel Hill - UL1TR002489: North Carolina Translational and Clinical Science Institute, University of Minnesota - UL1TR002494: Clinical and Translational Science Institute, Children's Hospital Colorado - UL1TR002535: Colorado Clinical and Translational Sciences Institute, The University of Iowa - UL1TR002537: Institute for Clinical and Translational Science, The University of Utah - UL1TR002538: Uhealth Center for Clinical and Translational Science, Tufts Medical Center - UL1TR002544: Tufts Clinical and Translational Science Institute, Duke University - UL1TR002553: Duke Clinical and Translational Science Institute, Virginia Commonwealth University - UL1TR002649: C. Kenneth and Dianne Wright Center for Clinical and Translational Research, The Ohio State University - UL1TR002733: Center for Clinical and Translational Science, The University of Miami Leonard M. Miller School of Medicine - UL1TR002736: University of Miami Clinical and Translational Science Institute, University of Virginia - UL1TR003015: iTHRIVL Integrated Translational health Research Institute of Virginia, Carilion Clinic - UL1TR003015: iTHRIVL Integrated Translational health Research Institute of Virginia, University of Alabama at Birmingham - UL1TR003096: Center for Clinical and Translational Science, Johns Hopkins University - UL1TR003098: Johns Hopkins Institute for Clinical and Translational Research, University of Arkansas for Medical Sciences - UL1TR003107: UAMS Translational Research Institute, Nemours - U54GM104941: Delaware CTR ACCEL Program, University Medical Center New Orleans - U54GM104940: Louisiana Clinical and Translational Science (LA CaTS) Center, University of Colorado Denver, Anschutz Medical Campus - UL1TR002535: Colorado Clinical and Translational Sciences Institute, Mayo Clinic Rochester - UL1TR002377: Mayo Clinic Center for Clinical and Translational Science (CCaTS), Tulane University - UL1TR003096: Center for Clinical and Translational Science, Loyola University Medical Center - UL1TR002389: The Institute for Translational Medicine (ITM), Advocate Health Care Network - UL1TR002389: The Institute for Translational Medicine (ITM), OCHIN - INV-018455: Bill and Melinda Gates Foundation grant to Sage Bionetworks
Additional data partners who have signed DTA and data release pending: The Rockefeller University - UL1TR001866: Center for Clinical and Translational Science, The Scripps Research Institute - UL1TR002550: Scripps Research Translational Institute, University of Texas Health Science Center at San Antonio - UL1TR002645: Institute for Integration of Medicine and Science, The University of Texas Health Science Center at Houston - UL1TR003167: Center for Clinical and Translational Sciences (CCTS), NorthShore University HealthSystem - UL1TR002389: The Institute for Translational Medicine (ITM), Yale New Haven Hospital - UL1TR001863: Yale Center for Clinical Investigation, Emory University - UL1TR002378: Georgia Clinical and Translational Science Alliance, Weill Medical College of Cornell University - UL1TR002384: Weill Cornell Medicine Clinical and Translational Science Center, Montefiore Medical Center - UL1TR002556: Institute for Clinical and Translational Research at Einstein and Montefiore, Medical College of Wisconsin - UL1TR001436: Clinical and Translational Science Institute of Southeast Wisconsin, University of New Mexico Health Sciences Center - UL1TR001449: University of New Mexico Clinical and Translational Science Center, George Washington University - UL1TR001876: Clinical and Translational Science Institute at Children's National (CTSA-CN), Stanford University - UL1TR003142: Spectrum: The Stanford Center for Clinical and Translational Research and Education, Regenstrief Institute - UL1TR002529: Indiana Clinical and Translational Science Institute, Cincinnati Children's Hospital Medical Center - UL1TR001425: Center for Clinical and Translational Science and Training, Boston University Medical Campus - UL1TR001430: Boston University Clinical and Translational Science Institute, The State University of New York at Buffalo - UL1TR001412: Clinical and Translational Science Institute, Aurora Health Care - UL1TR002373: Wisconsin Network For Health Research, Brown University - U54GM115677: Advance Clinical Translational Research (Advance-CTR), Rutgers, The State University of New Jersey - UL1TR003017: New Jersey Alliance for Clinical and Translational Science, Loyola University Chicago - UL1TR002389: The Institute for Translational Medicine (ITM), New York University - UL1TR001445: Langone Health's Clinical and Translational Science Institute, Children's Hospital of Philadelphia - UL1TR001878: Institute for Translational Medicine and Therapeutics, University of Kansas Medical Center - UL1TR002366: Frontiers: University of Kansas Clinical and Translational Science Institute, Massachusetts General Brigham - UL1TR002541: Harvard Catalyst, Icahn School of Medicine at Mount Sinai - UL1TR001433: ConduITS Institute for Translational Sciences, Ochsner Medical Center - U54GM104940: Louisiana Clinical and Translational Science (LA CaTS) Center, HonorHealth - None (Voluntary), University of California, Irvine - UL1TR001414: The UC Irvine Institute for Clinical and Translational Science (ICTS), University of California, San Diego - UL1TR001442: Altman Clinical and Translational Research Institute, University of California, Davis - UL1TR001860: UCDavis Health Clinical and Translational Science Center, University of California, San Francisco - UL1TR001872: UCSF Clinical and Translational Science Institute, University of California, Los Angeles - UL1TR001881: UCLA Clinical Translational Science Institute, University of Vermont - U54GM115516: Northern New England Clinical & Translational Research (NNE-CTR) Network, Arkansas Children's Hospital - UL1TR003107: UAMS Translational Research Institute

https://covid.cd2h.org/enclave-checklist

## Funding

The analyses described in this publication were conducted with data or tools accessed through the NCATS N3C Data Enclave covid.cd2h.org/enclave and supported by NCATS U24 TR002306. This research was possible because of the patients whose information is included within the data from participating organizations (covid.cd2h.org/dtas) and the organizations and scientists (covid.cd2h.org/duas) who have contributed to the on-going development of this community resource^2^.

The N3C data transfer to NCATS is performed under a Johns Hopkins University Reliance Protocol # IRB00249128 or individual site agreements with NIH. The N3C Data Enclave is managed under the authority of the NIH; information can be found at https://ncats.nih.gov/n3c/resources.

We gratefully acknowledge contributions from the following N3C core teams (leads noted with ^*^)

- Principal Investigators: Melissa A. Haendel^*^, Christopher G. Chute^*^, Kenneth R. Gersing, Anita Walden
- Workstream, subgroup and administrative leaders: Melissa A. Haendel^*^, Tellen D. Bennett, Christopher G. Chute, David A. Eichmann, Justin Guinney, Warren A. Kibbe, Hongfang Liu, Philip R.O. Payne, Emily R. Pfaff, Peter N. Robinson, Joel H. Saltz, Heidi Spratt, Justin Starren, Christine Suver, Adam B. Wilcox, Andrew E. Williams, Chunlei Wu
- Key liaisons at data partner sites
- Regulatory staff at data partner sites
- Individuals at the sites who are responsible for creating the datasets and submitting data to N3C
- Data Ingest and Harmonization Team: Christopher G. Chute*, Emily R. Pfaff^*^, Davera Gabriel, Stephanie S. Hong, Kristin Kostka, Harold P. Lehmann, Richard A. Moffitt, Michele Morris, Matvey B. Palchuk, Xiaohan Tanner Zhang, Richard L. Zhu
- Phenotype Team (Individuals who create the scripts that the sites use to submit their data, based on the COVID and Long COVID definitions): Emily R. Pfaff^*^, Benjamin Amor, Mark M. Bissell, Marshall Clark, Andrew T. Girvin, Stephanie S. Hong, Kristin Kostka, Adam M. Lee, Robert T. Miller, Michele Morris, Matvey B. Palchuk, Kellie M. Walters
- Project Management and Operations Team: Anita Walden^*^, Yooree Chae, Connor Cook, Alexandra Dest, Racquel R. Dietz, Thomas Dillon, Patricia A. Francis, Rafael Fuentes, Alexis Graves, Julie A. McMurry, Andrew J. Neumann, Shawn T. O’Neil, Usman Sheikh, Andréa M. Volz, Elizabeth Zampino
- Partners from NIH and other federal agencies: Christopher P. Austin^*^, Kenneth R. Gersing^*^, Samuel Bozzette, Mariam Deacy, Nicole Garbarini, Michael G. Kurilla, Sam G. Michael, Joni L. Rutter, Meredith Temple-O’Connor
- Analytics Team (Individuals who build the Enclave infrastructure, help create codesets, variables, and help Domain Teams and project teams with their datasets): Benjamin Amor^*^, Mark M. Bissell, Katie Rebecca Bradwell, Andrew T. Girvin, Amin Manna, Nabeel Qureshi
- Publication Committee Management Team: Mary Morrison Saltz^*^, Christine Suver^*^, Christopher G. Chute, Melissa A. Haendel, Julie A. McMurry, Andréa M. Volz, Anita Walden
- Publication Committee Review Team: Carolyn Bramante, Jeremy Richard Harper, Wenndy Hernandez, Farrukh M Koraishy, Federico Mariona, Saidulu Mattapally, Amit Saha, Satyanarayana Vedula
- Synthetic Data Domain Team including Yujuan Fu, Nisha Mathews, Ofer Mendelevitch

Data was provided from the following institutions: Stony Brook University — U24TR002306 • University of Oklahoma Health Sciences Center — U54GM104938: Oklahoma Clinical and Translational Science Institute (OCTSI) • West Virginia University — U54GM104942: West Virginia Clinical and Translational Science Institute (WVCTSI) • University of Mississippi Medical Center — U54GM115428: Mississippi Center for Clinical and Translational Research (CCTR) • University of Nebraska Medical Center U54GM115458: Great Plains IDeA-Clinical & Translational Research • Maine Medical Center — U54GM115516: Northern New England Clinical & Translational Research (NNE-CTR) Network • Wake Forest University Health Sciences — UL1TR001420: Wake Forest Clinical and Translational Science Institute Northwestern University at Chicago — UL1TR001422: Northwestern University Clinical and Translational Science Institute (NUCATS) • University of Cincinnati — UL1TR001425: Center for Clinical and Translational Science and Training • The University of Texas Medical Branch at Galveston — UL1TR001439: The Institute for Translational Sciences • Medical University of South Carolina — UL1TR001450: South Carolina Clinical & Translational Research Institute (SCTR) • University of Massachusetts Medical School Worcester — UL1TR001453: The UMass Center for Clinical and Translational Science (UMCCTS) • University of Southern California — UL1TR001855: The Southern California Clinical and Translational Science Institute (SC CTSI) • Columbia University Irving Medical Center — UL1TR001873: Irving Institute for Clinical and Translational Research • George Washington Children’s Research Institute — UL1TR001876: Clinical and Translational Science Institute at Children’s National (CTSA-CN) • University of Kentucky — UL1TR001998: UK Center for Clinical and Translational Science • University of Rochester — UL1TR002001: UR Clinical & Translational Science Institute • University of Illinois at Chicago — UL1TR002003: UIC Center for Clinical and Translational Science • Penn State Health Milton S. Hershey Medical Center — UL1TR002014: Penn State Clinical and Translational Science Institute • The University of Michigan at Ann Arbor — UL1TR002240: Michigan Institute for Clinical and Health Research • Vanderbilt University Medical Center — UL1TR002243: Vanderbilt Institute for Clinical and Translational Research • University of Washington — UL1TR002319: Institute of Translational Health Sciences • Washington University in St. Louis — UL1TR002345: Institute of Clinical and Translational Sciences • Oregon Health & Science University — UL1TR002369: Oregon Clinical and Translational Research Institute • University of Wisconsin-Madison — UL1TR002373: UW Institute for Clinical and Translational Research • Rush University Medical Center — UL1TR002389: The Institute for Translational Medicine (ITM) • The University of Chicago — UL1TR002389: The Institute for Translational Medicine (ITM) • University of North Carolina at Chapel Hill — UL1TR002489: North Carolina Translational and Clinical Science Institute • University of Minnesota — UL1TR002494: Clinical and Translational Science Institute • Children’s Hospital Colorado — UL1TR002535: Colorado Clinical and Translational Sciences Institute • The University of Iowa — UL1TR002537: Institute for Clinical and Translational Science • The University of Utah — UL1TR002538: Uhealth Center for Clinical and Translational Science • Tufts Medical Center — UL1TR002544: Tufts Clinical and Translational Science Institute • Duke University — UL1TR002553: Duke Clinical and Translational Science Institute • Virginia Commonwealth University — UL1TR002649: C. Kenneth and Dianne Wright Center for Clinical and Translational Research • The Ohio State University — UL1TR002733: Center for Clinical and Translational Science • The University of Miami Leonard M. Miller School of Medicine — UL1TR002736: University of Miami Clinical and Translational Science Institute • University of Virginia — UL1TR003015: iTHRIVL Integrated Translational health Research Institute of Virginia • Carilion Clinic — UL1TR003015: iTHRIVL Integrated Translational health Research Institute of Virginia • University of Alabama at Birmingham — UL1TR003096: Center for Clinical and Translational Science • Johns Hopkins University — UL1TR003098: Johns Hopkins Institute for Clinical and Translational Research • University of Arkansas for Medical Sciences — UL1TR003107: UAMS Translational Research Institute • Nemours — U54GM104941: Delaware CTR ACCEL Program • University Medical Center New Orleans — U54GM104940: Louisiana Clinical and Translational Science (LA CaTS) Center • University of Colorado Denver, Anschutz Medical Campus — UL1TR002535: Colorado Clinical and Translational Sciences Institute • Mayo Clinic Rochester — UL1TR002377: Mayo Clinic Center for Clinical and Translational Science (CCaTS) Tulane University — UL1TR003096: Center for Clinical and Translational Science • Loyola University Medical Center — UL1TR002389: The Institute for Translational Medicine (ITM) • Advocate Health Care Network — UL1TR002389: The Institute for Translational Medicine (ITM) • OCHIN — INV-018455: Bill and Melinda Gates Foundation grant to Sage Bionetworks

Additional data partners who have signed DTA and data release pending: The Rockefeller University — UL1TR001866: Center for Clinical and Translational Science • The Scripps Research Institute — UL1TR002550: Scripps Research Translational Institute • University of Texas Health Science Center at San Antonio — UL1TR002645: Institute for Integration of Medicine and Science • The University of Texas Health Science Center at Houston — UL1TR003167: Center for Clinical and Translational Sciences (CCTS) • NorthShore University HealthSystem — UL1TR002389: The Institute for Translational Medicine (ITM) • Yale New Haven Hospital — UL1TR001863: Yale Center for Clinical Investigation • Emory University — UL1TR002378: Georgia Clinical and Translational Science Alliance • Weill Medical College of Cornell University — UL1TR002384: Weill Cornell Medicine Clinical and Translational Science Center • Montefiore Medical Center — UL1TR002556: Institute for Clinical and Translational Research at Einstein and Montefiore • Medical College of Wisconsin — UL1TR001436: Clinical and Translational Science Institute of Southeast Wisconsin • University of New Mexico Health Sciences Center — UL1TR001449: University of New Mexico Clinical and Translational Science Center • George Washington University — UL1TR001876: Clinical and Translational Science Institute at Children’s National (CTSA-CN) • Stanford University — UL1TR003142: Spectrum: The Stanford Center for Clinical and Translational Research and Education • Regenstrief Institute — UL1TR002529: Indiana Clinical and Translational Science Institute • Cincinnati Children’s Hospital Medical Center — UL1TR001425: Center for Clinical and Translational Science and Training • Boston University Medical Campus — UL1TR001430: Boston University Clinical and Translational Science Institute The State University of New York at Buffalo — UL1TR001412: Clinical and Translational Science Institute Aurora Health Care — UL1TR002373: Wisconsin Network For Health Research • Brown University — U54GM115677: Advance Clinical Translational Research (Advance-CTR) • Rutgers, The State University of New Jersey — UL1TR003017: New Jersey Alliance for Clinical and Translational Science • Loyola University Chicago — UL1TR002389: The Institute for Translational Medicine (ITM) • New York University UL1TR001445: Langone Health’s Clinical and Translational Science Institute • Children’s Hospital of Philadelphia — UL1TR001878: Institute for Translational Medicine and Therapeutics • University of Kansas Medical Center — UL1TR002366: Frontiers: University of Kansas Clinical and Translational Science Institute Massachusetts General Brigham — UL1TR002541: Harvard Catalyst • Icahn School of Medicine at Mount Sinai — UL1TR001433: ConduITS Institute for Translational Sciences • Ochsner Medical Center — U54GM104940: Louisiana Clinical and Translational Science (LA CaTS) Center • HonorHealth — None (Voluntary) • University of California, Irvine — UL1TR001414: The UC Irvine Institute for Clinical and Translational Science (ICTS) • University of California, San Diego — UL1TR001442: Altman Clinical and Translational Research Institute • University of California, Davis — UL1TR001860: UCDavis Health Clinical and Translational Science Center • University of California, San Francisco — UL1TR001872: UCSF Clinical and Translational Science Institute • University of California, Los Angeles — UL1TR001881: UCLA Clinical Translational Science Institute • University of Vermont — U54GM115516: Northern New England Clinical & Translational Research (NNE-CTR) Network • Arkansas Children’s Hospital — UL1TR003107: UAMS Translational Research Institute Authorship was determined using ICMJE recommendations. The project described was supported by the National Institute of General Medical Sciences, 5U54GM104942-04. The content is solely the responsibility of the authors and does not necessarily represent the official views of the NIH. Any opinions expressed in this document are those of the authors and do not necessarily reflect the views of NCATS, individual N3C team members, or affiliated organizations and institutions.

### Contributor Statements

#### Masthead Authors

Authors ABW, JAT, NZ, RF, contributed to study conception and design. Author NZ contributed to the generation of the data. Author JAT conducted the experiment and data analysis. Author JAT wrote the manuscript with input from all authors. Authors ABW, RF led the N3C Synthetic Data Validation Task Team with support from author PP who led the broader N3C Synthetic Data Workstream.

#### Other Consortial Authors^*^

Christopher G. Chute^1,2,3,4,5,6,7,8,9,10^, Jon D. Morrow^1,12,13,2,7,9^, Melissa A. Haendel^14,6,10,11^

^1^clinical data model expertise, ^2^data curation, ^3^data integration, ^4^data quality assurance, ^5^funding acquisition, ^6^governance, ^7^critical revision of the manuscript for important intellectual content, ^8^N3C Phenotype definition, ^9^project evaluation, ^10^project management, ^11^regulatory oversight / admin, ^12^clinical subject matter expertise, ^13^data analysis, ^14^funding acquisition

^*^*Consortial authorship and corresponding contributions were self-reported as part of the N3C authorship committee review process*.

### Human Subjects Protections

This study was approved by the Washington University and University of Washington Internal Review Boards.

