## Supplemental table & figures for "Demonstrating an approach for evaluating synthetic geospatial and temporal epidemiologic data utility: Results from analyzing >1.8 million SARS-CoV-2 tests in the United States National COVID Cohort Collaborative (N3C)"

**SUPPLEMENT**

**Figure Titles:**

***S2:*** Distribution of total tests per zip code in original data which were censored within the synthetic data

***S3:*** MDClone data synthesis workflow

**Table S1:** Zip code month pairs' synthetic error central tendencies and counts stratified by indicator and bin size.

| **indicator** | **Number of zip codes stratified by month** | **Bin value original count** | **Synthetic Error mean (stdev)** | **Synthetic Error median (IQR)** |
|---|---|---|---|---|
| Tests | 33328 | 0-19 | -0.14 (±1.9) | 0 (2) |
| Tests | 5283 | 20-49 | -0.54 (±3.31) | -1 (5) |
| Tests | 2697 | 50-99 | -0.4 (±4.11) | 0 (5) |
| Tests | 2230 | 100-249 | -0.28 (±5.17) | 0 (6) |
| Tests | 1102 | 250-1705 | -0.59 (±7.29) | 0 (9) |
| Positives | 26707 | 0 | 0.07 (±0.37) | 0 (0) |
| Positives | 6499 | 1 | -0.55 (±0.92) | -1 (1) |
| Positives | 6264 | 2-5 | -0.78 (±1.76) | -1 (2) |
| Positives | 4715 | 6-49 | -0.59 (±2.63) | -1 (3) |
| Positives | 455 | 50-520 | -1.13 (±4.22) | -1 (5) |
| Admissions | 37963 | 0 | 0.04 (±0.25) | 0 (0) |
| Admissions | 3837 | 1 | -0.43 (±0.82) | -1 (1) |
| Admissions | 2078 | 2-4 | -0.66 (±1.42) | -1 (2) |
| Admissions | 499 | 5-9 | -1.37 (±2.29) | -1 (3) |
| Admissions | 263 | 10-80 | -2.16 (±3.33) | -2 (4) |

**Figure S2**

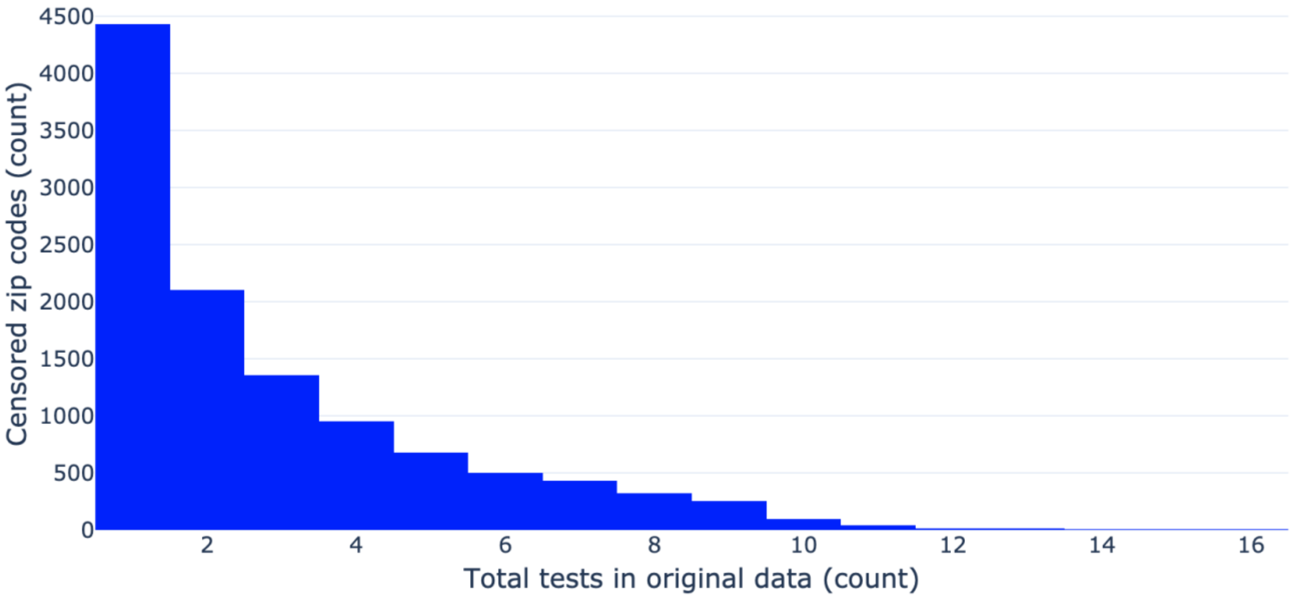


**Figure S3**

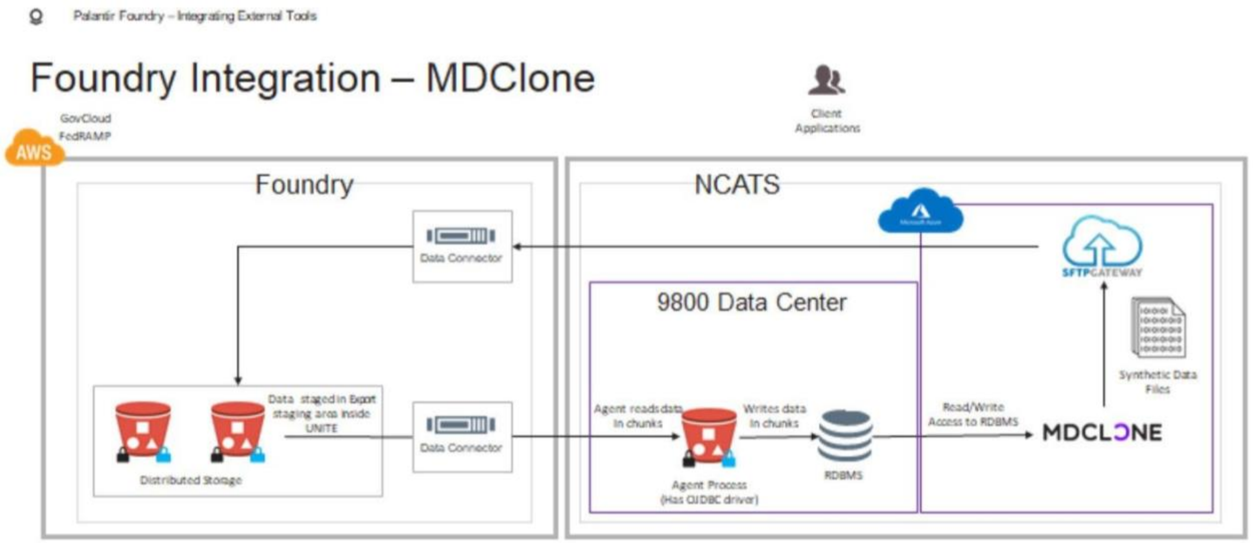
